# Glymphatic dysfunction in multiple sclerosis and its association with disease pathology and disability

**DOI:** 10.1101/2024.06.26.24309494

**Authors:** Ahmed Bayoumi, Khader M. Hasan, Akram Yazdani, John A. Lincoln

## Abstract

**Background:** MS is a chronic neuroinflammatory and neurodegenerative disease, in which the glymphatic system remains underexplored. The diffusion tensor image analysis along the perivascular space (ALPS) offers a non-invasive method to assess glymphatic function.

**Objective:** To investigate glymphatic function in MS, and its relationship with disease pathology and disability.

**Methods:** This retrospective study involved 118 MS patients, divided based on the Expanded Disability Status Scale (EDSS) into MS-L (EDSS < 3) and MS-H (EDSS ≥ 3). The ALPS index, brain parenchymal fraction (BPF), and lesion load (LL) were measured. Subgroup comparisons, ALPS index correlations with clinical and MRI measures, and logistic regression were performed.

**Results:** Significant differences in the mean ALPS index between MS-H and MS-L (d=-0.71, p-FDR=0.001) were found. The ALPS index correlated significantly with disease duration (rp=-0.29, p-FDR=0.002), and EDSS (rsp=-0.35, p-FDR=0.0002). The ALPS index also showed significant correlations with BPF and LL. The ALPS index and LL were significant predictors of lower disability (ALPS: Odds ratio (OR)=1.77, p=0.04, LL: OR=0.94, p=0.02).

**Conclusion:** Our findings highlight the ALPS index as an imaging biomarker in MS. This suggests the involvement of glymphatic dysfunction in MS pathology and underscores the need for further research to elucidate its role in MS.

## II Introduction

Multiple sclerosis (MS) is a chronic neuroimmunological disease characterized by central losses in myelin, oligodendrocytes, and axons. Current understanding of the underlying disease processes implicates elements from auto-reactive adaptive and innate immune systems, oxidative stress, cytotoxicity, as well as mitochondrial and blood-brain barrier dysfunction.^1^ Neuroinflammation and neurodegeneration are co-occurring features of MS pathology^2^, with studies highlighting the inception of neurodegeneration prior to the clinical onset of MS.^3^ Imaging measures have proved reliability in monitoring MS inflammatory and degenerative outcomes.^2, 4^ MS-related lesions have been long used as an imaging outcome for anti-inflammatory effects of MS treatments and predicted disability progression.^4^ Measurements of whole-brain volume, such as brain parenchymal fraction (BPF) have demonstrated sensitivity and reliability in quantifying neurodegeneration in MS.^5^

Cerebrospinal fluid (CSF) flow dynamics and their role in interstitial fluid (ISF) exchange have been proposed to play a role in various neuroinflammatory and neurodegenerative disease processes. The glymphatic system is purported to play a role in waste clearance in the central nervous system.^6^ First observed and proposed in animal models,^7^ the glymphatic system represents CSF influx from subarachnoid spaces via periarterial spaces into brain parenchyma facilitated by astroglial aquaporin-4 then into perivenous spaces, finally draining out of the brain into dural and cervical lymphatics.^8^

Advances in imaging have enabled the identification of several non-invasive surrogate measures for characterizing glymphatic function. Previously, this characterization was only possible through experiments using intrathecal,^9, 10^ or intravenous^11, 12^ tracer agents. A recent study employing Positron Emission Tomography with an intravenous tracer demonstrated CSF clearance deficits in PwMS.^12^ Additionally, the evaluation of perivascular spaces (PVS), also known as Virchow-Robin spaces, serves as another surrogate measure for glymphatic function.^13^ The literature denotes an increased burden of enlarged PVS in pwMS compared to healthy controls, albeit it shows heterogeneous associations with MS-related inflammatory and degenerative pathology.^14^ Recently developed methods have allowed for non-invasive evaluation of the glymphatic system using diffusion imaging, termed diffusion tensor image analysis along the perivascular space (DTI-ALPS).^15^ DTI-ALPS assesses water diffusion along perivascular space, leveraging the unique architecture at the level of lateral ventricles (LV), where medullary veins run perpendicular to the LV wall, neighboring projection fibers, and association fibers, allowing for specific assessment of diffusivity parallel to the medullary veins’ perivascular spaces.^15^

In this study, we aim to investigate the relationship between glymphatic system function and MRI markers for MS pathology, and how they associate with MS-related disability.

## III Methods

### Subjects

In this retrospective study, patients with MS (PwMS) diagnosed using the McDonald criteria at the comprehensive MS care clinic at UT Physicians, McGovern Medical School, UTHealth between 2011 and 2021, were included. Enrolled subjects underwent clinical examination, including the assessment of the Expanded Disability Status Scale (EDSS) score. Using a disability milestone EDSS score of 3.0 (moderate disability but no impairment of walking)^16^, the enrolled PwMS were divided into two subgroups. MS-L subgroup included PwMS with EDSS scores lower than 3, while MS-H subgroup included PwMS with EDSS scores greater than or equal to 3. The study was approved by the Institutional Review Board for the University of Texas Health Science Center at Houston – UTHealth. All participants in the study provided written consent following approved procedures by the Committee for the Protection of Human Subjects of McGovern Medical School, UTHealth.

### Image acquisition

MRI scans were performed using a 3.0 T Philips Ingenia research scanner with a maximum gradient amplitude of 45 mT/m and a 15-channel SENSE-compatible head coil (Philips Medical Systems, Best, Netherlands). High-resolution T^1^ magnetization prepared rapid gradient echo (MPRAGE) images (voxel size: 1 × 1 × 1 mm^3^, field of view (FOV): 256 × 256 mm^2^, TR/TE: 8/3.7 ms), used for anatomical registration, were acquired at the start of each scanning session, as well as T^2^-fluid-attenuated inversion recovery (FLAIR) images (voxel size: 1 × 1 × 1 mm^3^, FOV: 256 × 256 mm^2^, repetition time (TR)/ time to echo (TE): 4800/300 ms), which were used for lesion segmentation. Additionally, diffusion-tensor images (DTI) were obtained using a single-shot spin-echo diffusion-sensitized echo-planar imaging sequence with a balanced Icosa21 tensor encoding scheme.^17^ The b-factor was 1,000 s mm^−2^, TR/TE 7,100/65 ms, FOV = 256 × 256 mm, and the slice thickness was 3 mm with a 0 mm gap and a total of 44 slices. DTI quality control throughout data acquisition was assured using the same subject and water phantom tests as detailed previously.^18^ Further post-processing is described in these earlier works.^18, 19^

### Image analysis

#### A-DTI-ALPS processing

The DTI data were processed in FSL (version 6.0.6, FMRIB Software Library; http://www.fmrib.ox.ac.uk/fsl) as described by Taoka et al.^20^ DTI-ALPS evaluates water diffusion in the direction of the perivascular spaces, comparing the juxtaventricular projection and association diffusivities along the different three-dimensional axes. This area’s specific incongruent conformation between perivascular spaces, major projection, and association pathways allows for an approximate quantification of the perivascular space diffusivity.^15^ The DTI images underwent eddy current correction and tensor fitting using DTIFIT, obtaining the diffusivity maps in the x-axis (Dxx), y-axis (Dyy), and z-axis (Dzz) as well as the FA maps. The FA map of each subject was registered to the FMRIB58_FA standard space using linear and non-linear transformations, and obtained transformations were applied to the diffusivity maps. The subject with the least degree of warping was selected for region-of-interest (ROI) placement. Spherical ROIs with a diameter of 12 pixels were placed in the projection and association areas at the level of the LV body bilaterally confirmed on the color-coded FA map.

The ROIs were used to calculate the mean x-axis diffusivity in the projection (Dxxproj) and association (Dxxassoc) areas, the y-axis projection (Dyyproj) area diffusivity, and the z-axis association (Dzzassoc) area diffusivity. The ALPS index was calculated using the following formula:

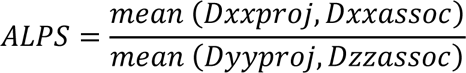

The ALPS indices of both hemispheres were calculated and averaged.

#### B-Structural image processing

In FSL, T_1_ and T_2_-FLAIR images were bias-field corrected and co-registered. In SPM12 (https://www.fil.ion.ucl.ac.uk/spm/software/spm12/), white matter (WM) hyperintense lesions were segmented using the Lesion Segmentation Toolbox^21^ and validated by a trained rater (AB) obtaining the lesion distribution map and the total lesion volumes for each subject. To mitigate the impact of MS lesions on spatial normalization,^22^ the produced lesion maps were used to perform lesion filling on the corresponding T_1_ images.^23^

Lesion-filled T_1_ images were processed in the Computational Anatomy Toolbox (CAT12; https://neuro-jena.github.io/cat), which has been validated in cross-sectional MS studies and shown to provide robust volumetric measures.^22, 24^ The T_1_ images were first denoised and affine-registered, after which they underwent unified segmentation^25^ which was further refined by applying a partial volume estimation^26^, finally they were spatially normalized using geodesic shooting registrations.^27^ Grey matter (GM), total WM, and CSF volumes were obtained as well as the total intracranial volume (TIV) and used to calculate brain parenchymal fraction (BPF = GM+WM / TIV), grey matter fraction (GMF = GM/TIV) and total WM fraction (WMF = WM/TIV). On the Neuromorphometrics atlas (Neuromorphometrics, Inc.), the tissue volumes for deep grey matter (DGM) structures and LV were estimated.^28^ To account for differences in TIV, lesion, DGM, and LV volumes were divided by TIV and multiplied by 1000, which were eventually used as the lesion load (LL), DGM, and LV volumes in the statistical analysis.

Cortical thickness was estimated and reconstructed using a projection-based method^29^ and topological defects were repaired using spherical harmonics.^30^ The created surfaces were mapped to the FreeSurfer “FsAverage” template^31^ to which the local thickness values were transferred and the mean cortical thickness (CTh) was obtained for each subject.

### Statistical analysis

In JASP (version 0.18.3, https://jasp-stats.org/), with a significance level set at *p* < 0.05, the demographic and clinical variables were compared between the subgroups MS-L and MS-H using *t*-test for continuous variables, Mann-Whitney U test for ordinal variables and Chi-squared test for categorical variables. Group-wise differences for the MRI measures were estimated using ANCOVA, controlling for age and gender. Post-hoc comparisons were performed to determine the mean differences, effect sizes as quantified by Cohen’s d (*d*), and 95% confidence intervals (CI) calculated from 5000 bootstraps.

The left and right ALPS index measurements were compared using a Paired Samples *t*-test and Pearson’s correlation coefficient between the mean ALPS index and age was calculated. To investigate the relationship between the ALPS index and the clinical and structural MRI measures, Pearson’s partial correlation coefficients (*r_p_*) were calculated for the continuous variables while Spearman’s rank correlation (*r_sp_*) was used to evaluate correlation with ordinal variables, adjusting for age and gender. P-values (*p*) from group differences and correlations were corrected for false discovery rate using the Benjamini-Hochberg procedure (*p*-FDR).

Multivariable logistic regression was performed to study the effects of the imaging measures and their association with the disability subgroups, MS-L and MS-H. In GraphPad Prism (version 10, GraphPad Inc., San Diego, CA, USA), we used the disability subgroup as a dependent variable and MS-L as the reference level, and BPF, LL, ALPS index, disease duration, age, and gender as covariates. The standardized regression coefficients (β) and odds ratios (OR) with 95% CI were calculated to assess the relationship between imaging variables and the disability subgroups, Wald’s test (*W*) was used to evaluate the significance of individual coefficients in the model. Area under the curve (AUC) was calculated using the receiver operating characteristic (ROC) curve, to assess the model’s overall performance. Individual ROC analyses were conducted, using the disability subgroup as the dependent variable, to assess the predictive power of the ALPS index, BPF, and LL in distinguishing between patients with lower and higher disability.

Raw data was generated at UTHealth. Anonymized derived data may be shared upon reasonable request to the corresponding or senior authors with researchers who provide a methodologically sound non-commercial proposal, subject to restrictions according to participant consent and data protection legislation.

## IV Results

A total of 118 PwMS were included in this study (mean age 45 ± 11.8; 65.3% female), divided into two subgroups. MS-L (n = 57; mean age 43.2 ± 12.1; 50.9% female) with EDSS < 3, and MS-H (n = 61; mean age 46.7 ± 11.4; 78.7% female) with EDSS ≥ 3. Their demographic and clinical data are summarized in Table 1.

**Table 1.** Demographic and clinical characteristics of PwMS with lower disability (MS-L) and higher disability (MS-H) Subject means for the whole group are first provided (MS), then divided into lower disability (MS-L) and higher disability (MS-H) groups for comparisons. Continuous variables were compared using student’s *t*-tests, categorical variables were assessed using Chi-squared tests, and EDSS was compared using Mann-Whitney U test. MS-L: lower disability subgroup; MS-H: higher disability subgroup; SD: standard deviation; EDSS: Expanded Disability Status Scale; IQR: inter-quartile range.

|  | **MS (n = 118)** | **MS-L (n = 57)** | **MS-H (n = 61)** | ***p*-value** |
| --- | --- | --- | --- | --- |
| **Age, mean (SD)** | 45 (11.8) | 43.2 (12.1) | 46.7 (11.4) | 0.1 |
| **Gender (% female)** | 65.3% | 50.9% | 78.7% | 0.002 |
| **Handedness (% right-handed)** | 90.7% | 94.7% | 86.9% | 0.1 |
| **Disease duration, mean (SD)** | 12.2 (8.5) | 10.6 (8.1) | 13.6 (8.6) | 0.1 |
| **EDSS, median (IQR)** | 3 (4) | 2 (1) | 6 (2.5) | < 0.001 |

### Imaging measures group differences

Compared to the MS-L group, the MS-H group exhibited significantly lower BPF (*d* = -0.42 [- 0.81, -0.03], *p*-FDR = 0.04) (Fig. 1A) and GMF (*d* = -0.41 [-0.79, -0.02], *p*-FDR = 0.04). The groups did not have significant differences in total WMF. Related to GM, MS-H showed prominent differences in DGM volumes (*d* = -0.7 [-1.09, -0.30], *p*-FDR = 0.001) and mean cortical thickness (*d* = -0.54 [-0.94, -0.15], *p*-FDR = 0.01). Lesion loads were significantly higher in MS-H compared to MS-L (*d* = 0.79 [0.39, 1.18], *p*-FDR = 0.001) (Fig. 1B). When examining the ALPS indices, we observed significant differences in mean ALPS index (*d* = - 0.71 [-1.11, -0.31], *p*-FDR = 0.001) (Fig. 1C). Table 2 summarizes group, subgroup and their differences adjusting for age and gender.

**Figure 1.**
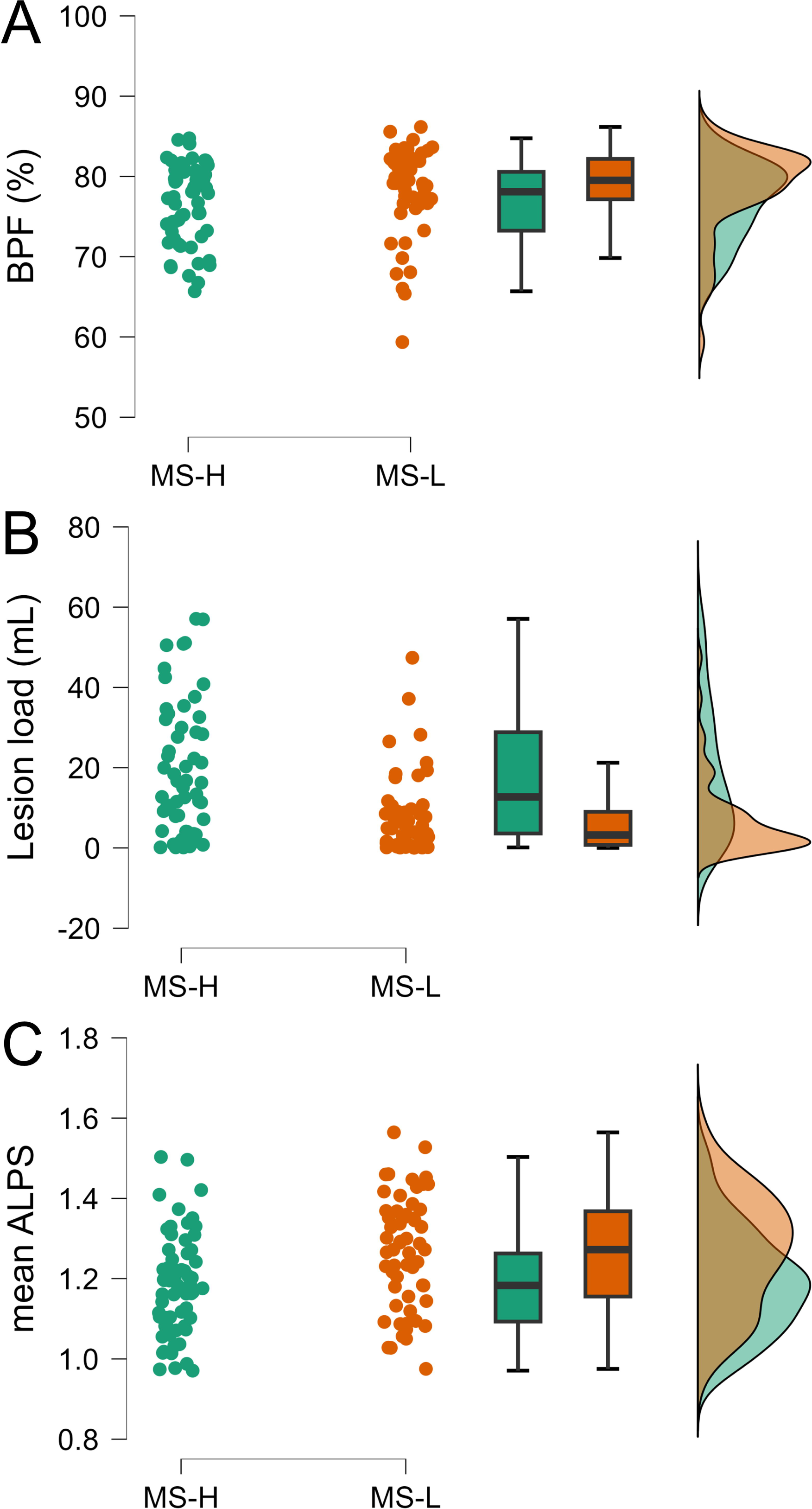
Group differences in brain parenchymal fraction, lesion load and mean ALPS index. Raincloud plots depicting the distribution and differences between the higher (MS-H, green) and the lower (MS-L, orange) disability subgroups. Differences were calculated using ANCOVA adjusting for age and gender. (A) Brain parenchymal fraction (BPF), (B) Lesion load, and (C) Mean ALPS.

**Table 2.** Group comparisons between pwMS with lower disability (MS-L) and higher disability (MS-H) for MRI measures: structural volumetric measurements and ALPS indices. Subject means for the whole group are first provided (MS, n =118), then divided into lower disability (MS-L, n = 57) and higher disability (MS-H, n = 61) groups for comparisons. MS-L: lower disability group; MS-H: higher disability group; *p*-FDR: false discovery rate adjusted p-value; GMF: grey matter fraction; WMF: white matter fraction; BPF: brain parenchymal fraction; CTh: cortical thickness; DGM: deep grey matter; LV: lateral ventricle; ALPS: diffusivity along perivascular spaces index.

|  | **MS** | **MS-L** | **MS-H** | **Effect size (*d*)** | ***p*-FDR** |
| --- | --- | --- | --- | --- | --- |
| **GMF (%)** | 41.2 (3.5) | 41.9 (3.2) | 40.5 (3.6) | - 0.41 (- 0.79, - 0.02) | 0.04 |
| **Total WMF (%)** | 36.5 (3.5) | 36.7 (3.5) | 36.3 (3.4) | - 0.19 (- 0.57, 0.20) | 0.3 |
| **BPF (%)** | 77.7 (5.2) | 78.6 (5.4) | 76.8 (4.8) | - 0.42 (- 0.81, - 0.03) | 0.04 |
| **CTh (mm)** | 2.17 (0.18) | 2.22 (0.16) | 2.12 (0.18) | - 0.54 (- 0.94, - 0.15) | 0.01 |
| **DGM (mL)** | 18.8 (3.1) | 19.8 (2.7) | 17.9 (3.3) | - 0.7 (- 1.09, - 0.30) | 0.001 |
| **LV (mL)** | 8.3 (4.2) | 7.3 (3.5) | 9.2 (4.6) | 0.47 (- 0.08, 0.86) | 0.03 |
| **Lesion load (mL)** | 12.8 (14.6) | 7.1 (9.7) | 18.1 (16.3) | 0.79 (0.39, 1.18) | 0.001 |
| **L-ALPS** | 1.25 (0.16) | 1.30 (0.16) | 1.20 (0.14) | - 0.74 (- 1.14, - 0.35) | 0.001 |
| **R-ALPS** | 1.20 (0.14) | 1.24 (0.14) | 1.17 (0.13) | - 0.57 (- 0.96, - 0.18) | 0.008 |
| **Mean ALPS** | 1.22 (0.14) | 1.27 (0.14) | 1.18 (0.13) | - 0.71 (- 1.11, - 0.31) | 0.001 |

### Correlations with DTI-ALPS

Overall, the left ALPS indices were higher than the right indices (*d* = 0.45 [0.26, 0.64], *p*-FDR = 0.003) (Fig. 2A) and the mean ALPS index showed a significant negative correlation with age (*r* = -0.27 [-0.46, -0.09], *p*-FDR = 0.003) (Fig. 2B). Significant negative correlations were found with disease duration (*r_p_* = -0.29 [-0.44, -0.12], *p*-FDR = 0.002) (Fig. 3A) and EDSS (*r_sp_* = -0.35 [-0.48, -0.19], *p*-FDR = 0.0002) (Fig. 3B).

**Figure 2.**
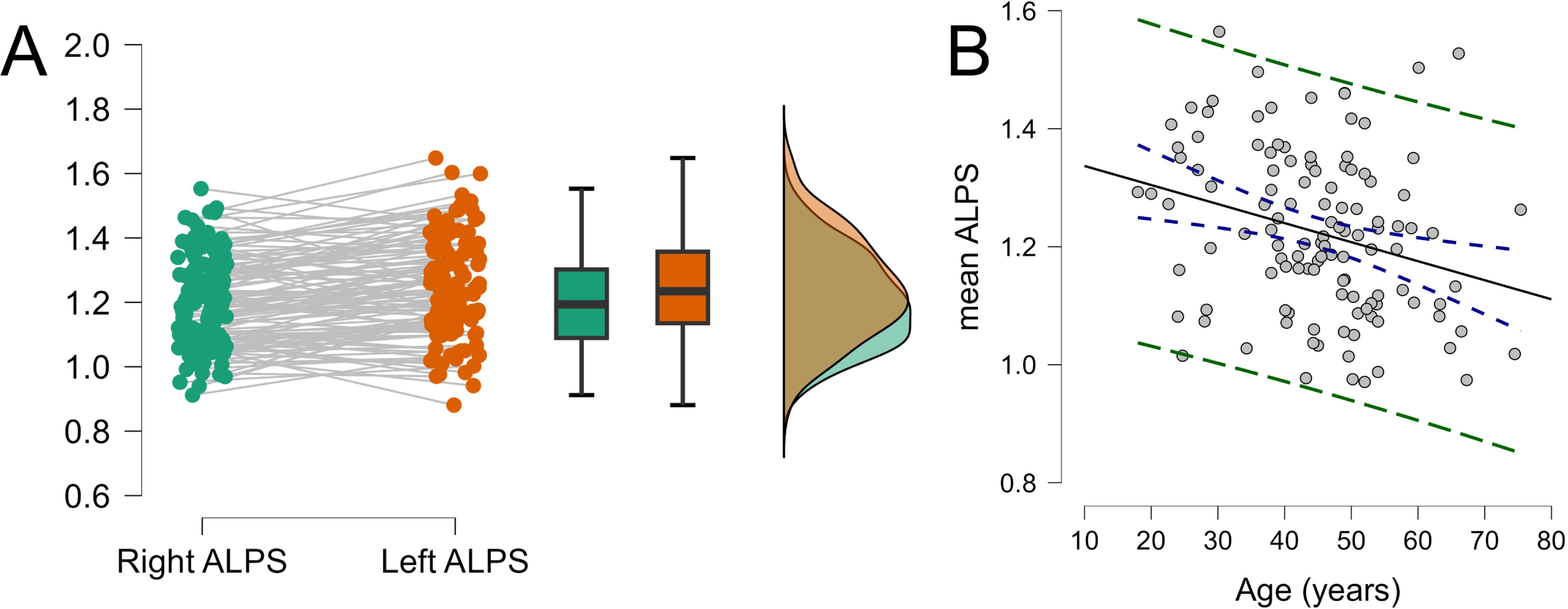
Paired differences between left and right ALPS indices and correlation between mean ALPS index and age. (A) Right and left ALPS index measurements differences estimated using paired samples *t*-test, (B) Correlation between mean ALPS index and age, blue dashes represent confidence interval and green dashes represent prediction intervals.

**Figure 3.**
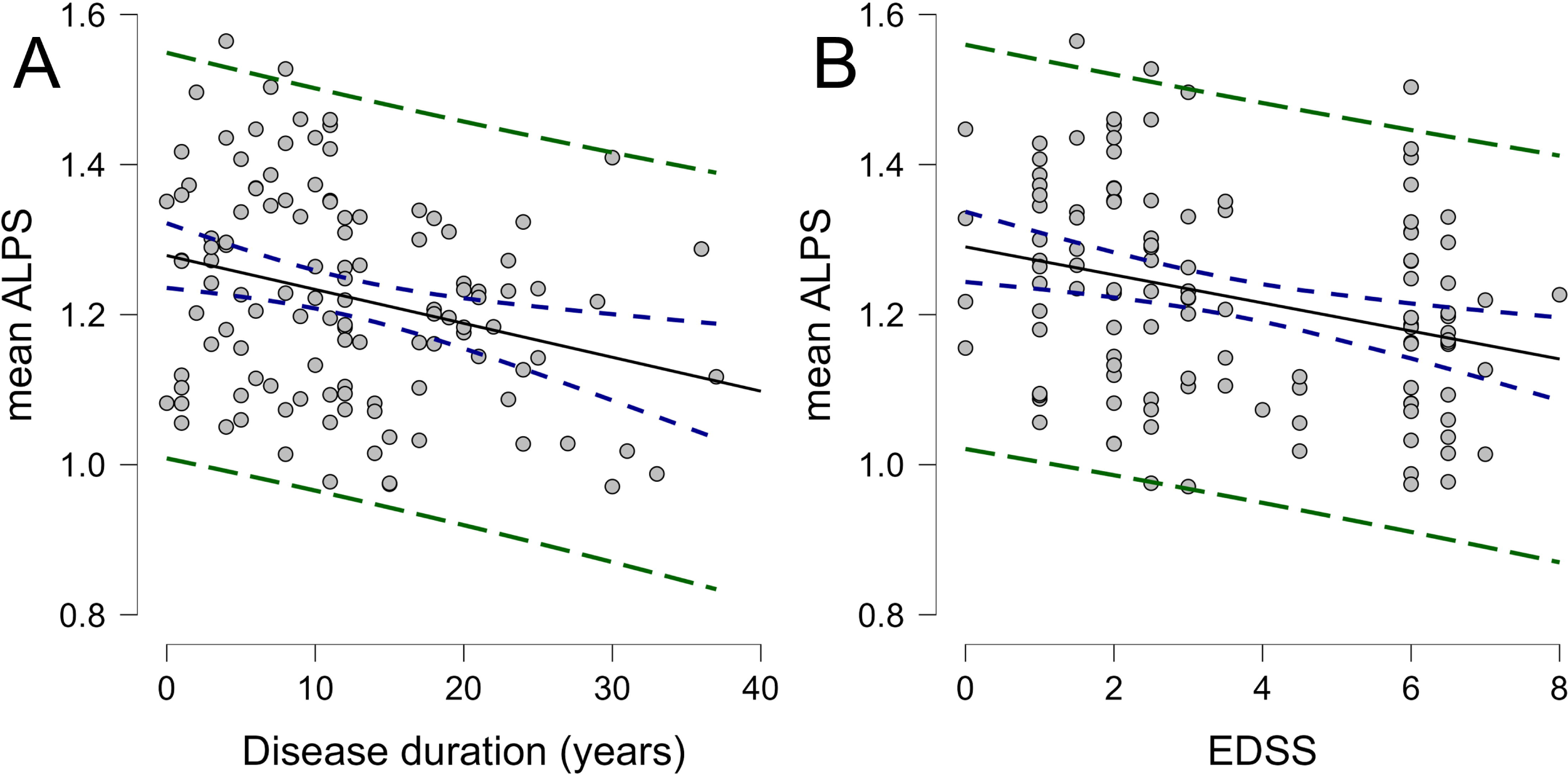
Correlations with disease duration and EDSS. (A) Pearson’s correlation calculated between mean ALPS index and disease duration adjusting for gender, (B) Spearman’s correlation calculated between ALPS index and EDSS adjusting for age and gender. Blue dashes represent confidence interval and green dashes represent prediction intervals.

For the imaging measures, ALPS index correlated positively with BPF (*r_p_* = 0.46 [0.31, 0.59], *p*-FDR < 0.0001) (Fig. 4A). Significant correlations were also observed with GMF (*r_p_* = 0.35 [0.18, 0.50], *p*-FDR = 0.0002) (Fig. 4B), DGM (*r_p_* = 0.44 [0.28, 0.57], *p*-FDR < 0.0001) (Fig. 4C), and CTh (*r_p_* = 0.29 [0.11, 0.45], *p*-FDR = 0.002) (Fig. 4D). In the WM, positive correlations were noted with total WMF (*r_p_* = 0.31 [0.15, 0.47], *p*-FDR = 0.0009) (Fig. 4E) and negative correlations with lesion load (*r_p_* = -0.49 [-0.62, -0.35], *p*-FDR < 0.0001) (Fig. 4F) (Table 3).

**Figure 4.**
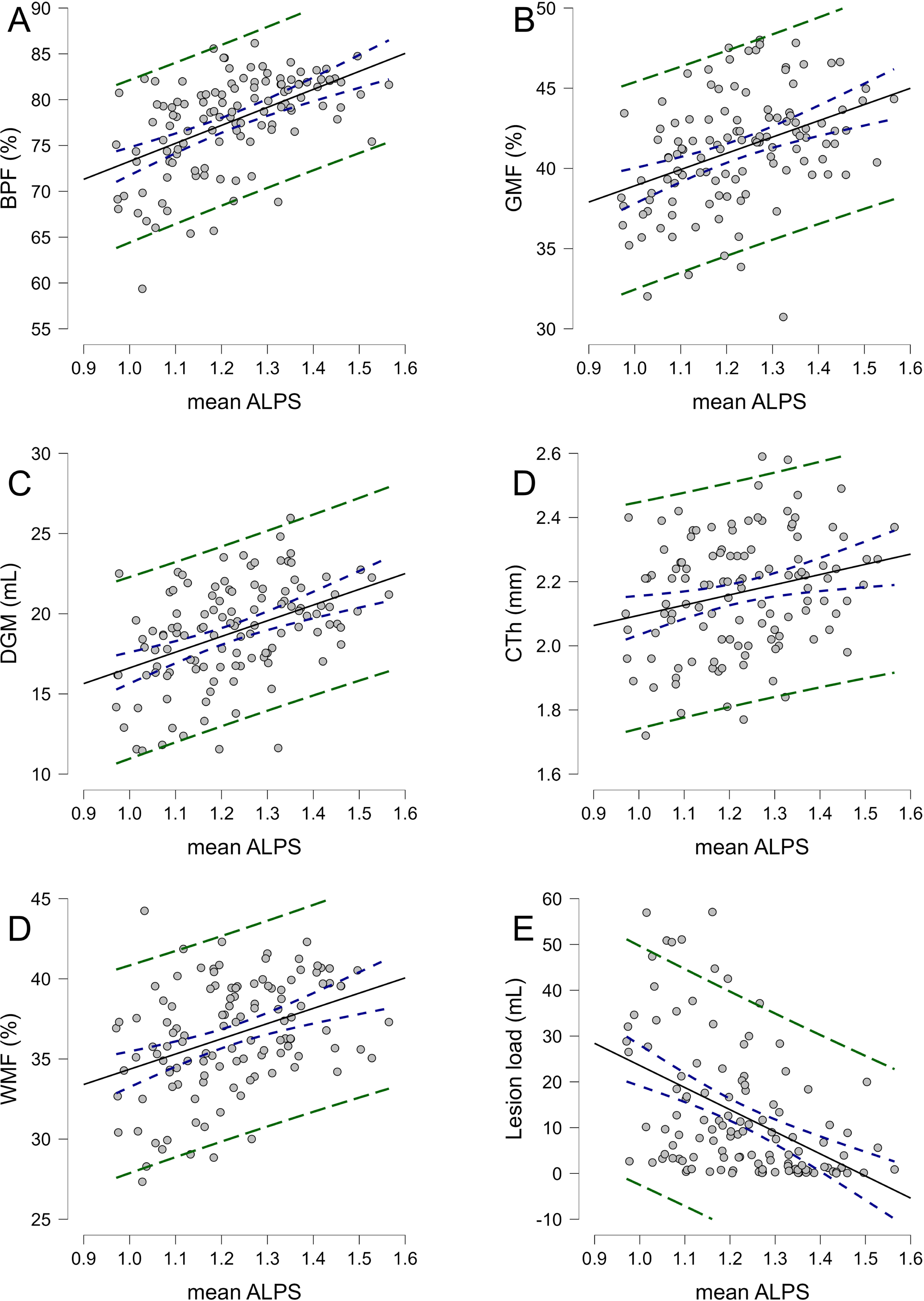
Correlations with imaging measures. Pearson’s partial correlations adjusting for age and gender between ALPS index and (A) Brain parenchymal fraction (BPF), (B) Grey matter fraction (GMF), (C) Deep grey matter (DGM), (D) Mean cortical thickness (CTh), (E) White matter fraction (WMF), and (F) Lesion load. Blue dashes represent confidence interval and green dashes represent prediction intervals.

**Table 3.**
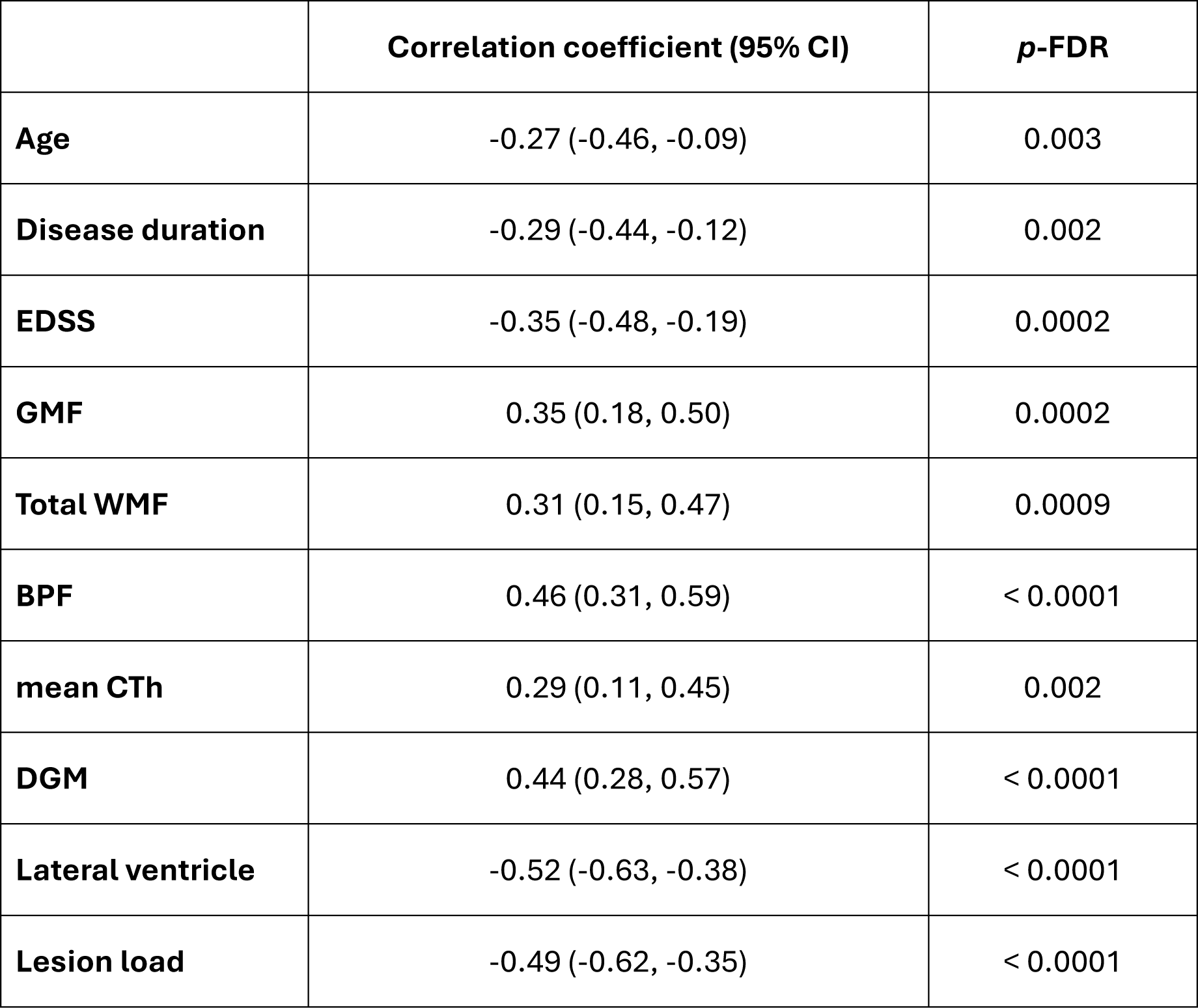
Correlations between mean ALPS index, disease duration, EDSS and volumetric MRI measures. CI: confidence interval; *p*-FDR: false discovery rate adjusted p-value; EDSS: Expanded Disability Status Scale; GMF: grey matter fraction; WMF: white matter fraction; BPF: brain parenchymal fraction; CTh: cortical thickness; DGM: deep grey matter; LV: lateral ventricle.

### DTI-ALPS discriminative power versus other imaging measures

The results of the multivariable logistic regression (Table 4), focusing on the relationship of disability subgroup with BPF, LL, and mean ALPS index, are summarized in Table 3. The model was significantly improved compared to the intercept-only model (Log-likelihood ratio (*G^2^*) = 33.41, *p* < 0.0001; AUC = 0.78 [0.70 – 0.86], *p* < 0.0001) (Fig. 5). The ALPS index was positively associated with lower disability (OR = 1.77 [1.05, 3.08], *W* = 4.39, *p* = 0.04), meanwhile, LL was positively associated with higher disability (OR = 0.94 [0.89, 0.98], *W* = 5.92, *p* = 0.01), and BPF was not a significant predictor (OR = 0.95 [0.84, 1.07], *W* = 0.77, *p* = 0.38).

**Figure 5.**
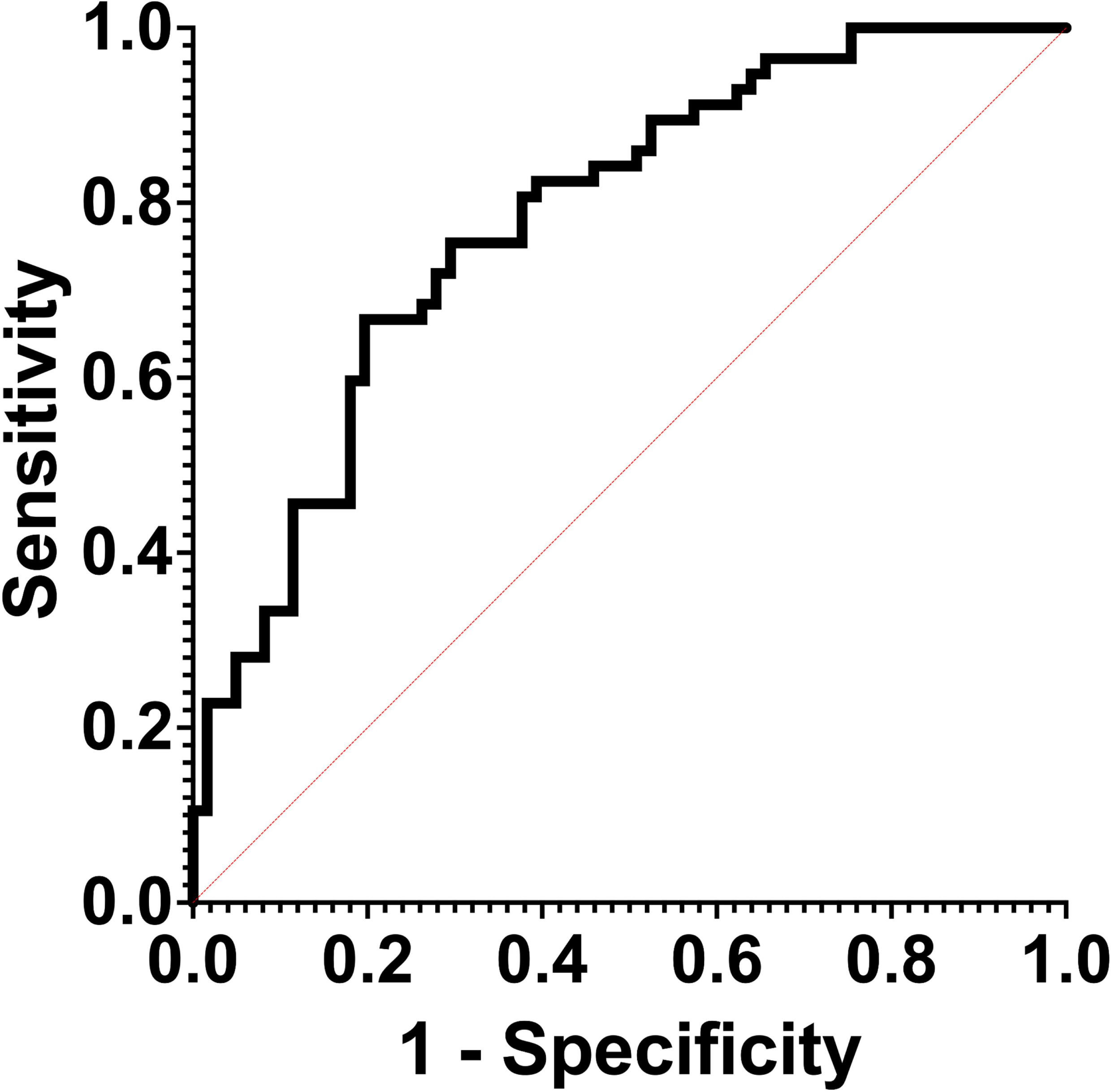
Multivariable logistic regression receiver-operating curve. ROC curve assessing the combined performance of the mean ALPS index, brain parenchymal fraction and lesion load in predicting disability level.

**Table 4.** Multiple logistic regression parameters showing association between imaging measures and disability subgroups. β: standardized regression coefficient; OR: odds ratio; CI: confidence interval; BPF: brain parenchymal fraction; ALPS: diffusion tensor image analysis along the perivascular space index.

| **Variable** | **$\beta$** | **OR (95% CI)** | **Wald Statistic (W)** | ***p*-value** |
| --- | --- | --- | --- | --- |
| **Age** | -0.20249 | 0.983 (0.94, 1.03) | 0.59 | 0.44 |
| **Gender (female)** | -0.63517 | 4.037 (1.57, 11.24) | 0.87 | 0.35 |
| **Disease duration** | 0.05164 | 1.006 (0.95, 1.07) | 0.04 | 0.83 |
| **BPF %** | -0.27887 | 0.9475 (0.84, 1.07) | 0.77 | 0.38 |
| **Lesion load** | -0.91428 | 0.9391 (0.89, 0.98) | 5.92 | 0.02 |
| **ALPS index** | 0.57023 | 1.769 (1.05, 3.08) | 4.39 | 0.04 |

Finally, individual ROC analyses were conducted for the ALPS index, LL, and BPF represented in Table 5 and Fig. 6. In summary, the largest AUC was observed with LL (AUC = 0.73, *p* < 0.0001), then the ALPS index (AUC = 0.67, *p* = 0.001), followed by BPF (AUC = 0.63, *p* = 0.01).

**Figure 6.**
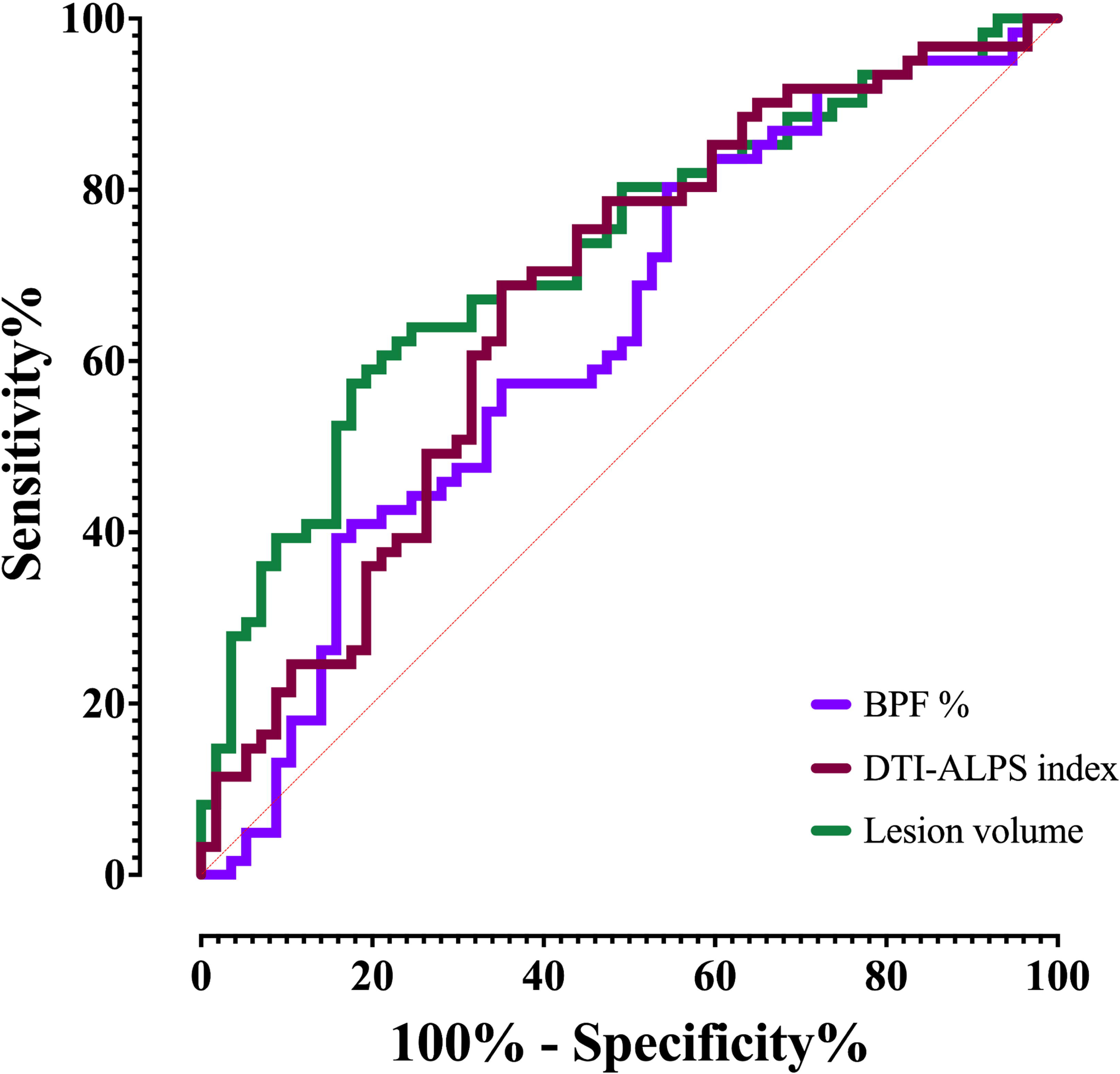
Independent receiver-operating curves for the mean ALPS index, brain parenchymal fraction and lesion load. ROC curves assessing the performance of each imaging measure in predicting disability level.

**Table 5.** Receiver-operating curve analysis results. AUC: areas under the curve; CI: confidence interval; ALPS: Diffusion tensor image analysis along the perivascular space index; BPF: brain parenchymal fraction.

|  | **AUC (95% CI)** | ***p*-value** |
| --- | --- | --- |
| **ALPS index** | 0.67 (0.57, 0.77) | 0.001 |
| **Lesion load** | 0.73 (0.63, 0.82) | < 0.0001 |
| **BPF** | 0.63 (0.53, 0.73) | 0.01 |

## V Discussion

In this study, we investigated glymphatic dysfunction and how it relates to MS-related disability and structural imaging MS correlates including brain atrophy measures and lesion load. Our results showed significant differences in the different imaging measures between the lower and higher disability subgroups. The most prominent decreases in MS-H compared to MS-L were observed in the ALPS indices, as well as deep grey matter volumes. MS-H had notable increases in lesion load compared to MS-L. When assessing the whole group for correlations between the mean ALPS index and various clinical and imaging measures, we found significant correlations with disease duration, EDSS, as well as grey matter fraction, total white matter fraction, and mean cortical thickness. Meanwhile, stronger correlations were found between the ALPS index and lesion load, brain parenchymal fraction, and deep grey matter. Using multiple logistic regression and ROC curves, we identified the ALPS index and lesion load to be significant parameters in distinguishing between MS-L and MS-H.

Previous studies have investigated glymphatic function in MS using the ALPS index,^32, 33^ Carotenuto et al. reported lower ALPS indices in pwMS (n = 71) compared to HC (n = 32), and PPMS (n = 22) compared to RRMS (n = 49).^32^ Another study assessed glymphatic function in MS by comparing HC (n = 13), patients with RRMS (n=13), and patients with SPMS (n =13) showing lower ALPS indices in the pwMS compared to HC, and in SPMS compared to RRMS patients. The authors reported a lack of significant correlation with disease duration.^33^

Our study focused on investigating the interplay between glymphatic function, MS-related imaging measures, and disability. Our lower and higher disability subgroups had significant differences in ALPS indices, despite not showing significant differences in disease duration. Moreover, ALPS index and lesion load, but not BPF, could predict reaching the EDSS 3.0 milestone in our sample, which might allude to a more fundamental role than association with neurodegeneration or brain atrophy.

Our study has a few limitations. First, its retrospective cross-sectional design limits inferences on the role of glymphatic dysfunction in the evolution of MS pathology regarding causality and temporality. This highlights the need for longitudinal studies to better characterize the role of glymphatic function during MS emergence and evolution. Second, our study lacks healthy controls to act as a referent group, although we aimed to investigate the relationship between glymphatic function, MS-related imaging, and disability outcomes. Third, we refrained from categorizing the subjects according to their respective MS phenotypes, given the retrospective nature of the study and the bias it could have introduced. To that point, there is growing evidence suggesting that clinical phenotypes may be more of a continuum rather than pathologically distinct conditions.^34^

## VI Conclusion

This study elucidates the relationship between glymphatic dysfunction, as quantified by the ALPS index, MS pathology, and disability. Our results demonstrate that decreased ALPS indices correlate with higher disability, increased lesion loads, and increased brain atrophy, suggesting a possible role of glymphatic dysfunction in MS progression. The ALPS index emerges as a possible imaging biomarker, distinguishing between different levels of MS-related disability and correlating with clinical and imaging measures. Despite its limitations, including a cross-sectional design and the absence of healthy controls, this study highlights the potential value of further research on glymphatic function in MS, urging further exploration in longitudinal studies to better understand its role in the disease process.

## VIII Data availability statement

Raw data was generated at UTHealth. Anonymized derived data may be shared upon reasonable request to the corresponding or senior authors with researchers who provide a methodologically sound non-commercial proposal, subject to restrictions according to participant consent and data protection legislation.

